# Non-Communicable Diseases Risk Factors and the Risk of COVID-19 Infection among University Employees in Indonesia

**DOI:** 10.1101/2022.01.17.22269249

**Authors:** Indah Suci Widyahening, Dhanasari Vidiawati, Trevino A Pakasi, Pradana Soewondo, Abdillah Ahsan

## Abstract

**Introduction:** Non-communicable diseases (NCDs) are still a major public health problem in Indonesia. Studies have shown that risk factors of NCDs were associated with coronavirus disease 2019 (COVID-19) severity and mortality. However, it is unclear whether NCD risk factors are also risks for new COVID-19 infection. This study aimed to obtain the NCD risk profiles among university’s employees and its association with COVID-19 infection.

**Methods:** A cross-sectional study was conducted in October 2021. Participants were administrative employees of Universitas Indonesia (UI), Depok City, West Java. Assessment of NCDs risk factors was based on the World Health Organization STEPwise approach to NCD risk factor surveillance (WHO STEPS). Demographics, working and medical history data were obtained electronically by using a Google Form. Physical and laboratory examination were done in the Integrated Post for NCDs. Risks were expressed as adjusted odds ratio (OR_adj_) and 95% confidence interval (CI) in multivariate analyses.

**Results:** A total of 613 employees were enrolled. Men were predominant (54.8%) and about 36% of them work in shift as security personnel. About 66.7% were overweight or obese and 77.8% had hypertension. There were 138 (22.8%) employees who had COVID-19 infection. Nearly all (95.6%) have completed COVID-19 vaccination. At-risk waist circumference (OR_adj_ 1.74, 95% CI 1.17-2.60, p=0.007) and total cholesterol level of 200-239 mg/dL (OR_adj_ 2.24, 95% CI 1.15-4.33, p=0.017) were independent risk factors, but shift work (OR_adj_ 0.54, 95% CI 0.24-0.84, p=0.006) was protective to COVID-19 infection.

**Conclusion:** The prevalence of NCDs risk factors among university administrative employees was high, increasing the risk for COVID-19 infection. A behavioral intervention program to manage the NCD risk factors at the university level is urgently needed according to the Health Promoting University framework.

## INTRODUCTION

The coronavirus disease 2019 (COVID-19) pandemic caused by the severe acute respiratory syndrome coronavirus 2 (SARS-CoV-2) is still ongoing and possibly become endemic. Currently, community transmission of COVID-19 in Indonesia has remained at low level after a second dramatic peak in mid-July 2021.[1] However, the actual number of cases may be higher than the officially registered; the weighted estimate of seroprevalence of SARS-CoV-2 antibody in Jakarta in March 2021 was 44.5%. This implies that almost half of the population has been infected with SARS-CoV-2.[2]

Non-communicable diseases (NCDs) are major health problems in the world. The national Basic Health Research in 2018 has shown increasing prevalence of NCDs in Indonesia, such as diabetes (from 6.9% in 2013 to 10.9% in 2018) and hypertension (from 25.8% in 2013 to 34.1% in 2018).[3] During the early COVID-19 pandemic, patients needing intensive care unit were more likely to have NCDs as comorbidities (hypertension, diabetes, cardiovascular disease, and cerebrovascular disease).[4]

The association of NCD risk factors and COVID-19 infection is not clear. Previous data showed that body mass index (BMI) and obesity are associated with COVID-19 infection, hospitalization and mortality.[5,6] Obesity seems play an important role in the pathogenesis of COVID-19.[7] Multimorbidity, especially renal, cardiovascular and metabolic morbidities, is associated with a higher risk of COVID-19 positive test.[8,9]

To prevent new infections in the future, we need to identify the modifiable risk factors for COVID-19, such as obesity and/or components of metabolic syndrome. This is important because factors that can be modified to reduce risk will be the target for health education along with rigorous vaccination program. This study aimed to obtain the NCD risk profiles among university’s employees and its association with COVID-19 infection.

## METHODS

### Study design and participants

The design of this study was cross-sectional and conducted in October 2021. Participants were employees of the University Central Administration, Universitas Indonesia (UI) and the administrative/supporting staffs of the Faculty Medicine of the university. Demographics, working and medical history data were obtained electronically by using a Google Form, which was distributed through the WhatsApp group of the Directorate of Human Resources and the Faculty of Medicine. Participants were then invited to attend physical and laboratory examination in the Integrated Post for NCDs throughout Universitas Indonesia at main campus at Depok City, West Java and the Salemba campus at Jakarta (total 6 posts). This post was a community-based program oriented towards promotive and preventive efforts to control NCDs.[10]

Sample size was calculated using the formula for a prevalence study with the confidence level of 0.05 and a precision of 5%. Hence a minimal number of 384 subjects is expected. Participants were enrolled in this study if they agree to give written consent. Ethics approval was granted from the Health Research Ethic Committee of the Faculty of Medicine Universitas Indonesia – Cipto Mangunkusumo Hospital (No. KET-1006/UN2.F1/ETIK/PPM.00.02/2021). Written informed consent was requested from all participants prior to the study.

### History of COVID-19 infection and vaccination

Diagnosis of COVID-19 was established by a history of positive PCR or rapid detection test for SARS-CoV-2 from nasopharyngeal swabs. However, participants might choose not to reveal their test results. History of COVID-19 infection was counted once between March 2020 and October 2021; re-infection, if occurred, was not considered. Participants were also asked whether or not they were hospitalized from COVID-19.

COVID-19 vaccination was started in January 2021 (in UI Hospital) and in mid-2021, most non-medical university employees have been scheduled for two doses.

### Assessment of non-communicable diseases risk factors

Assessment was based on the World Health Organization STEPwise approach to NCD risk factor surveillance (STEPS).[11] The survey instrument includes: tobacco use, alcohol use, physical inactivity, unhealthy diet, and key biological risk factors: overweight and obesity, raised blood pressure, raised blood glucose, and abnormal blood lipids. Risk factors assessed in this study consisted of anthropometric measurements (body mass index and waist circumference), medical history (hypertension, diabetes, dyslipidemia), working pattern (shift or non-shift), behavioral risk factors (smoking, alcohol drinking, fruit, vegetable, and salt consumption, and physical activity), and blood chemistry results (fasting blood glucose; triglyceride; total, low-density lipoprotein (LDL), and high-density lipoprotein (HDL) cholesterol levels.

Body weight was measured using a weight scale (SECA, Hamburg, Germany), and body height was measured by using a microtoise. The BMI was calculated as body weight in kilogram (kg) divided by body height in meter squared. Nutritional status was then classified based on the BMI criteria for Asian population as follows: underweight (BMI <18.5 kg/m^2^), normal weight (18.5–22.9 kg/m^2^), overweight (23.0–24.9 kg/m^2^), obesity class I (25.0–29.9 kg/m^2^), and obesity class II (>30.0 kg/m^2^).[12] Waist circumference (WC) was measured using a measuring type. At-risk WC was >102 cm for men and >88 cm for women.

Systolic and diastolic blood pressures (BP) were measured using a digital blood pressure monitor. Hypertension was indicated if systolic BP was ≥130 mmHg and/or diastolic BP ≥85 mmHg. A minimum of 8 hours fasting was required for the measurement of fasting blood glucose (FBG), triglyceride (TG) and cholesterol levels. The result was indicated as high if FBG ≥110 mg/dL, TG ≥ 150 mg/dL, total cholesterol ≥200 mg/dL, LDL-cholesterol ≥100 mg/dL, while HDL-cholesterol was considered low if ≤40 mg/dL. Physical activity was considered low (inactive) if less than 10 minutes and less than 2 times per week.

### Statistical analyses

Baseline characteristics of the participants and risk factors were presented descriptively. Univariate logistic regression test was performed for each risk factor. Multivariate logistic regression analyses were done to estimate the adjusted odds ratio (OR_adj_) and its corresponding 95% confidence intervals (CIs) for the association between risk factor and COVID-19 infection. A p-value of less than 0.05 was considered significant. All statistical analyses were done using Statistical Package for Social Science (SPSS) version 20.

## RESULTS

### Characteristics of the study subjects

From 789 supporting staffs invited, 750 attended the examination, 618 completed the questionnaire and 5 did not undergo laboratory tests. Totally, 613 (77.7%) data were finally included for analyses. Table 1 shows the characteristics and health history of the university employees. Male participants were predominant (54.8%). Around 79.7% of them were 40 years old or less and 72.4% were married. About 36% (222 of 613) work in shift as security personnel. There were 138 (22.8%) employees who had a history of positive COVID-19 but only 14 (2.3%) who need to be hospitalized. Nearly all of the participants (95.6%) have completed COVID-19 vaccination.

**Table 1.** Characteristics and health history of the university employees (n=613)

| Variables | n (%) |
| --- | --- |
| Age; mean (SD): 34.81 (15.10) |  |
| ≤ 40 years | 483 (79.7) |
| > 40 years | 123 (20.3) |
| Sex |  |
| Male | 336 (54,8) |
| Female | 277 (45,2) |
| Marriage status |  |
| Married | 444 (72.4) |
| Un-married | 169 (27.6) |
| Working in shift |  |
| Yes | 222 (36.2) |
| No | 391 (63.8) |
| History of hypertension |  |
| Yes | 143 (23.6) |
| No | 462 (75.4) |
| History of diabetes |  |
| Yes | 21 (3.4) |
| No | 584 (96.6) |
| History of dyslipidemia |  |
| Yes | 103 (16.8) |
| No | 501 (83.2) |
| History of COVID-19 infection |  |
| Yes | 138 (22.8) |
| No | 467 (77.2) |
| History of COVID 19 hospitalization |  |
| Yes | 14 (2.3) |
| No | 592 (97.7) |
| COVID 19 vaccination |  |
| Complete | 576 (95.6) |
| In-complete | 17 (2.8) |
| None | 10 (1.6) |

Table 2 shows the distribution of behavioral risk factors for NCDs among the university employees. Proportion of those who currently smoking is near to 25% (151 of 613) and 15% (94 of 613) admitted consuming alcohol. Proportion of those who consume fruit 5-7 days a week was just below 20% (120 of 613), while for vegetable consumption it is also not reaching 50% (285 of 613). Majority of the respondents (80%) are categorized as having an active physical activity level.

**Table 2.** Behavioral risk factors for non-communicable diseases among university employees (n=613)

| Variable | n (%) |
| --- | --- |
| Current smoking |  |
| Yes | 151 (24.6) |
| No | 462 (75.4) |
| Alcohol consumption |  |
| Yes | 94 (15.2) |
| No | 524 (84.8) |
| Fruit consumption |  |
| Never | 12 (2.0) |
| 1-4 days/week | 483 (78.5) |
| 5-7 days/week | 120 (19.5) |
| Vegetable consumption |  |
| Never | 5 (0.8) |
| 1-4 days/week | 325 (52.8) |
| 5-7 days/week | 286 (46.4) |
| Salt consumption |  |
| Far too much or too much | 114 (18.5) |
| Just the right amount | 314 (51.0) |
| Too little or far too little | 114 (18.5) |
| Don't know | 74 (12.0) |
| Physical activity level |  |
| Inactive | 119 (19.4) |
| Active | 494 (80.6) |

Table 3 shows biological risk factors for NCDs among the university employees. Two third of them (409 of 613) are categorized as overweight and obese, more than 75% (477 of 613) have an increased blood pressure, half (320 of 613) are having an at-risk waist circumference and almost half (294 of 613) are having an increased fasting blood sugar.

**Table 3.**
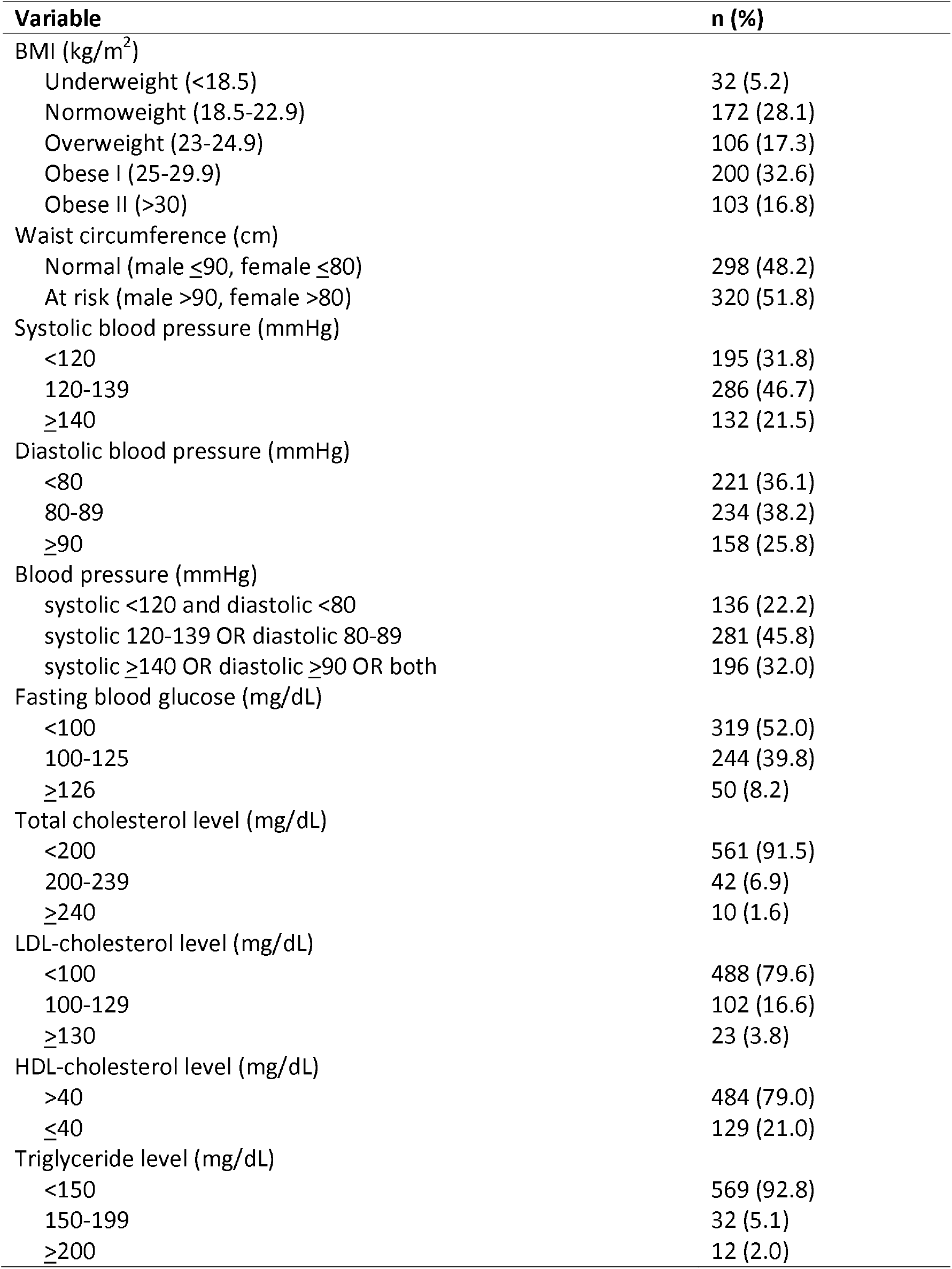
Biological risk factors for non-communicable diseases among university employees (n=613)

Table 4 shows variables that have moderate relationship (p<0.2) during bivariate analysis with the history of COVID 19 infection. Shift work (Adjusted OR [OR_adj_] 0.54, 95% Confidence Interval [CI] 0.24-0.84, p=0.006) having at risk waist circumference (OR_adj_ 1.74, 95% CI 1.17-2.60, p=0.007), and total cholesterol level of 200-239 mg/dl (OR_adj_ 2.24, 95% CI 1.15-4.33, p=0.017) are variables that have significant relationship with history of COVID-19 infection during multivariate logistic regression.

**Table 4.** Risk factors of COVID 19 infection among university employees (n=613)

| Variables | OR | 95% CI | $p^*$ | $OR_{adj}$ | 95%CI | $p^\dagger$ |
| --- | --- | --- | --- | --- | --- | --- |
| Working in shift |  |  |  |  |  |  |
| No | Ref |  |  | Ref |  |  |
| Yes | 0.50 | 0.32-0.77 | **0.010** | 0.54 | 0.35-0.84 | **0.006** |
| Smoking |  |  |  |  |  |  |
| No | Ref |  |  | Ref |  |  |
| Yes | 0.51 | 0.31-0.84 | **0.007** | 0.701 | 0.40-1.23 | 0.214 |
| Fruit consumption |  |  |  |  |  |  |
| 5-7 days/week | Ref |  |  | Ref |  |  |
| 1-4 days/week | 0.67 | 0.43-1.06 | 0.086 | 0.82 | 0.51-1.32 | 0.408 |
| Never | 0.22 | 0.03-1.79 | 0.157 | 0.31 | 0.04-2.52 | 0.271 |
| Salt consumption |  |  |  |  |  |  |
| Just the right amount/too little/far too little | Ref |  |  | Ref |  |  |
| Far too much/too much/don't know | 0.63 | 0.83-0.99 | **0.037** | 0.68 | 0.43-1.06 | 0.088 |
| BMI ($kg/m^2$) | | | | | | |
| <23 | Ref |  |  | Ref |  |  |
| 23-24.9 | 1.19 | 0.66-2.16 | 0.559 | 0.94 | 0.49-1.80 | 0.856 |
| $\geq 25$ | 1.66 | 2.07-2.59 | **0.024** | 1.10 | 0.59-2.04 | 0.767 |
| Waist circumference (cm) |  |  |  |  |  |  |
| Normal (male $\leq 90$, female $\leq 80$) | Ref | | | Ref | | |
| At risk (male $> 90$, female $> 80$) | 1.81 | 1.23-2.68 | **0.003** | 1.74 | 1.17-2.60 | **0.007** |
| Total Cholesterol (mg/dl) |  |  |  |  |  |  |
| <200 | Ref |  |  | Ref |  |  |
| 200-239 | 2.54 | 1.33-4.86 | **0.005** | 2.24 | 1.15-4.33 | **0.017** |
| $\geq 240$ | 2.99 | 0.79-11.30 | 0.107 | 2.85 | 0.73-11.13 | 0.131 |
| LDL (mg/dl) |  |  |  |  |  |  |
| <100 | Ref |  |  | Ref |  |  |
| 100-129 | 1.17 | 0.70-1.94 | 0.547 | 0.77 | 0.41-1.45 | 0.425 |
| $\geq 130$ | 3.04 | 1.28-7.24 | **0.012** | 0.95 | 0.24-3.82 | 0.944 |

\* univariate logistic regression
† multivariate logistic regression

## DISCUSSION

Our study showed that more than half of the university employees have at least one biological risk factor of NCDs (overweight or obese, increased blood pressure, at-risk waist circumference, and increased fasting blood sugar). Having at-risk waist circumference and total cholesterol level of 200-239 mg/dL increased the risk of COVID 19 infection while working in shift prevented it.

Assessing the NCD risk factors was our university commitment to protect health and promotes the well-being of the university members as part of the Health Promoting University framework.[13] It seems that NCD risk factors are common among university employees. A study in Saudi also found that more than half of the university employees had 3 or more NCD risk, i.e., 64% were overweight or obese, 22.1% had hypertension, and 21.5% had diabetes.[14] Another study showed that 72% of the university employees and their families were overweight or obese.[15] In Nigeria, the most common risk factors among university employees were inadequate intake of fruit and vegetables (94.6%), physical inactivity (77.8%), and dyslipidemia (51.8%).[16]

In the early COVID-19 pandemic, obesity was identified as a significant risk factor for severe disease.[17,18] A population-based cohort study found that excess weight is an important modifiable risk factor for severe COVID-19 outcomes.[19] Meta-analyses confirmed that obesity is a risk factor for developing severe COVID-19 through several possible mechanisms.[20]

A large cohort study from a general population in UK found that modifiable risk factors for COVID-19 were higher BMI, higher glycated hemoglobin, smoking, slow walking pace (a proxy for physical fitness), and use of blood pressure medication (a proxy for hypertension). High level of HDL cholesterol was associated with lower risk. The authors then concluded that lifestyle modification might reduce the risk of COVID-19.[21] Further study found that higher total cholesterol and ApoB levels might increase the risk of COVID-19 infection.[22]

Many studies used BMI as an indicator of obesity, which means excessive body fat. However, higher BMI may not represent higher amount of body fat since it cannot distinguish between fat and lean body mass.[23] We found that high BMI did not have significant association with the incidence of COVID-19 but high waist circumference (WC) did. If we look at our participants, about 30% of them work as security guards, implying that they may have higher BMI due to higher muscle mass. Therefore, WC is a better indicator for excessive body fat since it directly indicates the abdominal or central obesity.

Clinical studies on abdominal obesity and COVID-19 are still evolving. A study found that abdominal obesity was associated with a high chest x-ray severity score better than BMI.[24] UK study found that high WC was associated with positive COVID-19 test only in people ≥ 65 years, independent of BMI.[25] Another study found a positive association between WC and COVID-19 susceptibility (OR□= □1.38; 95% CI: 1.07–1.78; *p*=0.015). However, the significance was lost after adjustment for BMI. The authors concluded that overall obesity has a causal impact on the susceptibility of COVID-19 and obese people are regarded as high-risk.[26]

The ACE2 receptor plays an important role for SARS-COV-2 infectivity as the virus use the receptor to attach to the cell surface.[27] ACE2 is widely expressed in fat and may be the reason why obese patients experience more severe COVID-19 patients.[28] A recent study found that the expression of ACE2 in fat tissue (both visceral and subcutaneous) is higher than in lung tissue.[29] Upon binding to the cell surface, SARS-CoV-2 may enter the cell through clathrin-mediated endocytosis of the cell membrane.[30] Human cell membrane is made from cholesterol, a key structural lipid that is often used by pathogens for their pathogenesis.[31] Therefore, higher membrane cholesterol provides higher efficiency of the virus entry. However, COVID-19 patients generally showed reduced total cholesterol, HDL, and LDL-cholesterol levels which were associated with disease severity.[32] Systematic review confirmed that lower total, HDL- and LDL-cholesterol levels were significantly associated with COVID-19 severity and mortality, but not triglyceride level.^33^

On the contrary of other studies, working in shift was a protective factor for COVID-19 infection. For instance, a study in UK found that shift work was associated with increased risk of COVID-19.[34] More detail analyses showed that shift work was associated with a higher likelihood of COVID-19 for both irregular (OR 2.42; 95% CI 1.92–3.05) and permanent shift work (OR 2.50; 95% CI 1.95–3.19).[35]

There were several limitations in this study. First, the design was not prospective and included only people who survived COVID-19. Secondly, we cannot differentiate whether the participants had COVID-19 infection before or after vaccination, and most of the employees completed their vaccination about three months before our data collection. Therefore, the effect of the COVID-19 vaccine on the incidence was not known. Universitas Indonesia is one of the most prominent universities in Indonesia, located in an area where the prevalence of NCDs and COVID-19 is the highest in the country. Hence the prevalence of COVID-19 infection and NCD risk factors reported in this study might also be higher than other university employees in Indonesia.

## CONCLUSION

University administrative employees have a substantially high prevalence of NCD risk factors and this has increased their risk for COVID-19 infection. A behavioral intervention program to manage the NCD risk factors at the university level is urgently needed according to the Health Promoting University framework.

## Data Availability

All relevant data are within the manuscript and its Supporting Information files

## ACKNOWLEDGEMENTS

The authors would like to express their gratitude to the Director of Human Resources of Universitas Indonesia and the Dean Office of the Faculty of Medicine Universitas Indonesia for their support during data collection. The authors also thank the Makara Satellite Clinic of Universitas Indonesia for their assistance in the physical and laboratory examination of the subjects. Likewise, the authors are thankful for the assistance of Firky Ditha Saputri, MD and Helisa Rachel, MD during data collection and analysis.

## References

1. World Health Organization. Indonesia Situation Report – 81. Available at: https://cdn.who.int/media/docs/default-source/searo/indonesia/covid19/external-situation-report-81_17-november-2021.pdf?sfvrsn=9edbe641_5.

2. Ariawan I, Jusril H, Farid MN, Riono P, Wahyuningsih W, Widyastuti, et al. SARS-CoV-2 antibody seroprevalence in Jakarta, Indonesia: March 2021. Available at SSRN: https://ssrn.com/abstract=3954041.

3. Ministry of Health (MoH), Republic of Indonesia. National Institute of Health Research and Development. Basic Health Research 2018. Jakarta: MoH, 2018.

4. Wang D, Hu B, Hu C, Zhu F, Liu X, Zhang J, et al. Clinical characteristics of 138 hospitalized patients with 2019 novel Coronavirus-infected pneumonia in Wuhan, China. JAMA. 2020;323:1061–9.

5. Tartof SY, Qian L, Hong V, Wei R, Nadjafi RF, Fischer H, et al. Obesity and mortality among patients diagnosed with COVID-19: results from an integrated health care organization. Ann Intern Med. 2020;173(10):773–81. https://doi.org/10.7326/M20-3742.

6. Hendren NS, de Lemos JA, Ayers C, Das SR, Rao A, Carter S, et al. Association of body mass index and age With morbidity and mortality in patients hospitalized with COVID-19: Results from the American Heart Association COVID-19 Cardiovascular Disease Registry. Circulation. 2021; 143(2):135–44. https://doi.org/10.1161/CIRCULATIONAHA.120.051936.

7. Kassir R. Risk of COVID-19 for patients with obesity. Obes Rev. 2020;21(6):e13034. https://doi.org/10.1111/obr.13034.

8. McQueenie R, Foster HME, Jani BD, et al. Multimorbidity, polypharmacy, and COVID-19 infection within the UK Biobank cohort. PLoS One 2020;15:e0238091.

9. Woolford SJ, D’Angelo S, Curtis EM, Parsons, CM, Ward KA, Dennison EM, et al. COVID-19 and associations with frailty and multimorbidity: a prospective analysis of UK Biobank participants. Aging Clin Exp Res 2020;32:1897–1905. doi:10.1007/s40520-020-01653-6.

10. Tri Siswati T, Margono, Husmarini N, Purnamaningrum YE, Bunga Astria Paramashanti BA. Health-promoting university: the implementation of an integrated guidance post for non-communicable diseases (Posbindu PTM) among university employees. Global Health Promotion. 2021 (accepted article). https://doi.org/10.1177/17579759211021363.

11. World Health Organization. STEPwise Approach to NCD Risk Factor Surveillance (STEPS). Available at: https://www.who.int/teams/noncommunicable-diseases/surveillance/systems-tools/steps.

12. World Health Organization. Regional Office for the Western Pacific. The Asia-Pacific perspective: redefining obesity and its treatment. Sydney, Australia: Health Communications Australia, 2000.

13. Mónica Suárez-Reyes M, Van den Broucke S. Implementing the Health Promoting University approach in culturally different contexts: a systematic review. Global Health Prom. 2016; 23 Supp. 1: 46–56.

14. Amin TT, Al Sultan AI, Mostafa OA, Darwish AA, Al-Naboli MR. Profile of Non-Communicable Disease Risk Factors Among Employees at a Saudi University. Asian Pac J Cancer Prev. 2014;15 (18):7897–7. http://dx.doi.org/10.7314/APJCP.2014.15.18.7897.

15. Alzeidan R, Rabiee F, Mandil A, Hersi A, Fayed A. Non-communicable disease risk factors among employees and their families of a Saudi university: An epidemiological study. PLoS ONE. 2016;11(11):e0165036. https://doi.org/10.1371/journal.pone.0165036.

16. Agaba EI, Akanbi MO, Agaba PA, Ocheke AN, Gimba ZM, Daniyam S, et al. A survey of non-communicable diseases and their risk factors among university employees: a single institutional study. Cardiovasc J Afr 2017; 28: 377–84.

17. Földi M, Farkas N, Kiss S, et al. Obesity is a risk factor for developing critical condition in COVID-19 patients: a systematic review and meta-analysis. Obes Rev 2020; 21: e13095.

18. Huang Y, Lu Y, Huang Y-M, et al. Obesity in patients with COVID-19: a systematic review and meta-analysis. Metabolism 2020; 113: 154378.

19. Gao M, Piernas C, Astbury NM, Hippisley-Cox J, O’Rahilly S, Aveyard P, et al. Associations between body-mass index and COVID-19 severity in 6.9 million people in England: a prospective, community-based, cohort study. Lancet Diabetes Endocrinol 2021; 9: 350–9. https://doi.org/10.1016/S2213-8587(21)00089-9.

20. Aghili SMM, Ebrahimpur M, Arjmand B, Shadman Z, Pejman Sani M, Qorbani M, et al. Obesity in COVID-19 era, implications for mechanisms, comorbidities, and prognosis: a review and meta-analysis. Int J Obes (Lond). 2021;45(5):998–1016. https://doi.org/10.1038/s41366-021-00776-8.

21. Ho FK, Celis-Morales CA, Gray SR, Katiireddi SV, Niedzwieds CL, Hastie C, et al. Modifiable and non-modifiable risk factors for COVID-19, and comparison to risk factors for influenza and pneumonia: results from a UK Biobank prospective cohort study. BMJ Open 2020;10(11):e040402. https://doi.org/10.1136/bmjopen-2020-040402.

22. Zhang K, Dong S-S, Guo Y, Tang S-H, Wu H, Yao S, et al. Causal associations between blood lipids and COVID-19 risk: A two-sample Mendelian randomization study. Arterioscl Thromb Vasc Biol. 2021;41:2802–10. https://doi.org/10.1161/ATVBAHA.121.316324

23. Okorodudu DO, Jumean MF, Montori VM, Romero-Corral A, Somers VK, Erwin PJ et al (2010) Diagnostic performance of body mass index to identify obesity as defined by body adiposity: a systematic review and meta-analysis. Int J Obes 34:791–799. https://doi.org/10.1038/ijo.2010.5.

24. Malavazos AE, Secchi F, Basilico S, Capitanio G, Boveri S, Milani V, et al. Abdominal obesity phenotype is associated with COVID-19 chest X-ray severity score better than BMI-based obesity. Eat Weight Disord. 2021. https://doi.org/10.1007/s40519-021-01173-w.

25. Christensen RAG, Sturrock SL, Arneja J, Brooks JD. Measures of adiposity and risk of testing positive for SARS-CoV-2 in the UK Biobank Study. J Obesity. 2021;2021:8837319. https://doi.org/10.1155/2021/8837319.

26. Freuer D, Linseisen J, Meisinger M. Impact of body composition on COVID-19 susceptibility and severity: A two-sample multivariable Mendelian randomization study. Metabolism. 2021;118:154732. https://doi.org/10.1016/j.metabol.2021.154732.

27. Hoffmann M, Kleine-Weber H, Schroeder S, Krüger N, Herrler T, Erichsen S, et al. SARS-CoV-2 cell entry depends on ACE2 and TMPRSS2 and is blocked by a clinically proven protease inhibitor. Cell. 2020;181:271–80.

28. Al-Benna S. Association of high level gene expression of ACE2 in adipose tissue with mortality of COVID-19 infection in obese patients. Obes Med. 2020;19:100283. https://doi.org/10.1016/j.obmed.2020.100283.

29. Jia X, Yin C, Lu S, Chen Y, Liu Q, Bai J, Lu Y. Two things about COVID-19 might need attention. Preprints 2020, 2020020315. https://doi.org/10.20944/preprints202002.0315.v1

30. Bayati A, Kumar R, Francis V, McPherson PS. SARS-CoV-2 infects cells after viral entry via clathrin-mediated endocytosis. J Biol Chem. 2021;296:100306. https://doi.org/10.1016/j.jbc.2021.100306.

31. Dang EV, Madhani HD, Vance RE. Cholesterol in quarantine. Nat Immunol. 2020; 21(7):716–7.

32. Kocar E, Rezen T, Rozman D. Cholesterol, lipoproteins, and COVID-19: Basic concepts and clinical applications. Biochim Biophys Acta Mol Cell Biol Lipids. 2021;1866:158849. https://doi.org/10.1016/j.bbalip.2020.158849.

33. Zinellu A, Panagiotis P, Fois AG, Solidoro P, Carru C, Mangoni AA. Cholesterol and triglyceride concentrations, COVID-19 severity, and mortality: A systematic review and meta-analysis with meta-regression. Front Public Health. 2021;9:1210. https://doi.org/10.3389/fpubh.2021.705916.

34. Fatima Y, Bucks RS, Mamun AA, Skinner I, Rosenzweig I, Leschziner G, et al. Shift work is associated with increased risk of COVID-19: Findings from the UK Biobank cohort. J Sleep Res. 2021;30:e13326. https://doi.org/10.1111/jsr.13326.

35. Maidstone R, Anderson SG, Ray DW, Rutter MK, Durrington HJ, Blaikley JF. Shift work is associated with positive COVID-19 status in hospitalised patients. Thorax 2021;76:601–6.

